# Pediatric critical care capacity in Canada: a national cross-sectional study

**DOI:** 10.1101/2022.12.07.22283061

**Authors:** Saptharishi Lalgudi Ganesan, Daniel Garros, Jennifer Foster, Tanya Di Genova, Patricia S. Fontela, Srinivas Murthy, the Canadian Critical Care Trials Group (CCCTG)

## Abstract

**Background:** Pediatric intensive care unit (PICU) capacity is a current and future health system challenge in Canada. Despite experiencing two pandemics over the last 15 years and surges in PICU admissions every winter, the bed capacity of Canadian PICUs and their ability to accommodate surges in demand are unknown.

**Methods:** We conducted an internet-based cross-sectional survey to gather information from Canadian PICUs regarding PICU characteristics, medical staffing, therapies provided, and anticipated challenges related to surge management. The survey was completed by a representative of each PICU and validated by PICU Directors. Quantitative survey results were summarized as counts, proportions, and ratios while qualitative response was analyzed using inductive content analysis.

**Results:** Representatives from all **19 PICUs** located in **17 hospitals** completed the survey and reported having **275** (**217** level 3 & **58** level 2) funded beds with **298** physical bed spaces. Two PICUs representing **47** beds (**35** Level 3 & **12** Level 2) are specialized cardiac ICUs. Roughly **13385, 13419, 11430 and 12315** Canadian children were admitted to these PICUs in the years 2018, 2019, 2020 & 2021, respectively. During a surge, PICUs reported being able to add **5.9 ± 3.4 (range: 0 – 14)** beds per unit and a total of **108** temporary surge beds. Several barriers for the successful implementation of surge plans were identified.

**Interpretation:** Canadian pediatric critical care capacity is comparable to other high-income countries, though our ability to respond to a pandemic/epidemic surge with significant pediatric critical illness may be limited.

## Introduction

Appropriate assessment and management of critically ill children requires a specialized multidisciplinary healthcare team and dedicated facilities (1, 2). Pediatric intensive care units (PICU) have an integral role in managing acute illness, post-operative care, trauma, and complications of chronic diseases in children. As such, intensive care resources are limited and costly (3). While a ‘*PICU bed*’ includes the actual bed space, with access to electrical outlets, medical gases, and equipment (both monitoring and therapeutic), in healthcare system administration, a ‘*funded PICU bed*’ includes the physical bed within the unique ecosystem of trained pediatric intensivists, critical care nurses, pediatric respiratory therapists, specialized pediatric consulting services, and other members of the interprofessional team (4, 5) (figure 1). PICU capacity must be considered in the context of specialized staffing availability.

**Figure 1:**
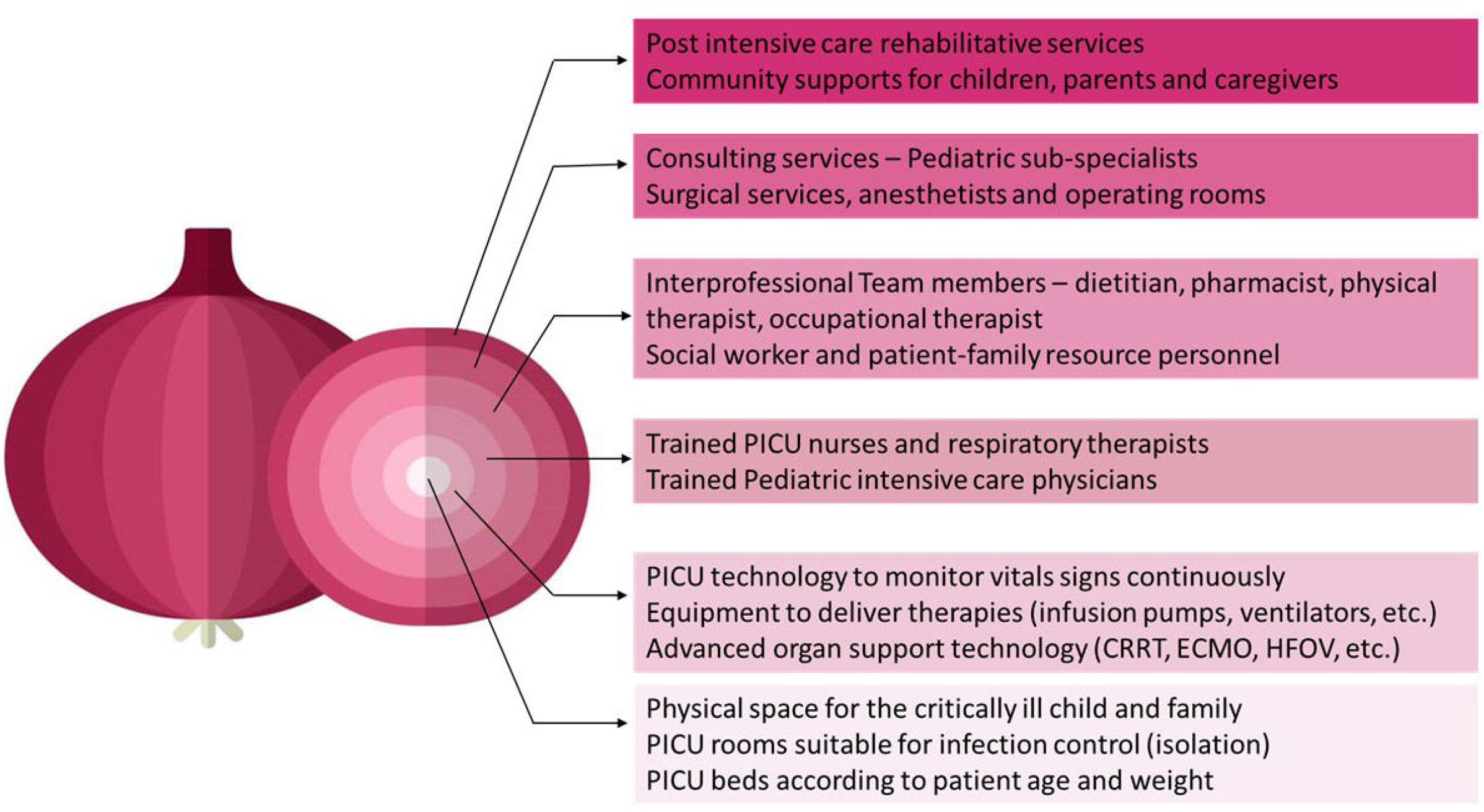
Onion peel model of a ‘funded’ pediatric intensive care unit bed Footnote: CRRT – Continuous renal replacement therapy, ECMO – Extra-corporeal membrane oxygenation, HFOV – High frequency oscillatory ventilation

Globally, the pandemic forced healthcare systems to evaluate their critical care capacity and ability to cope with capacity challenges (6–9). While adult ICUs bore the brunt of pandemic-related admissions, the comparatively smaller capacity of pediatric critical care in Canada was also strained (10). Despite few Canadian children with COVID-19 requiring PICU admission, pre-existing capacity limitations were accentuated, leading to strains in service delivery that continue post-pandemic (11).

The recent surge of viral infections (12) has caused a dramatic strain on capacity across Canada and exposed several gaps in the ability of country’s PICUs to meet demands during surges (13, 14). Despite these recent and ongoing challenges, Canadian national PICU capacity is unknown. Planning for ongoing and future surges in demand, growing and shifting population trends, and resource assignment requires an understanding of current PICU capacity. Therefore, the objectives of our cross-sectional study were to define the capacity of Canadian PICUs and understand their ability to accommodate surges in demand.

## Methods

### Study Design

We designed an internet-based cross-sectional survey to address the study objectives. The survey design, dissemination and reporting were carried out in line with the CROSS-reporting guidelines(15). The Office of Human Research Ethics (OHRE) determined that this study did not require oversight by Western University’s research ethics board.

### Data collection instruments

Physician members from 5 different Canadian PICUs developed a survey questionnaire to address the following domains: hospital characteristics, ICU characteristics, medical staffing, therapies provided, and surge management in the context of a pandemic. The study team participated in an item generation and reduction process to ensure assessment of each objective. The draft version of the questionnaire was uploaded into the SurveyMonkey® platform and piloted by 4 pediatric critical care physicians (MDs from 4 different provinces). The testers completed a sensibility questionnaire to provide feedback regarding the survey questionnaire’s clarity, relevance, completeness, face validity, content validity, redundancy, and time for completion. We refined the questionnaire based on the feedback received.

We considered a PICU to be one in which care for critically ill children is provided by a fellowship-trained pediatric critical care specialist, and a team (nursing, respiratory therapist) with pediatric critical care-specific skill sets. A priori, we agreed that it would not be possible to assign levels of care for PICUs based on definitions provided by the American Academy of Pediatrics and the Society of Critical Care Medicine, unlike in the neonatal intensive care context where levels are more clearly defined (16–18). For the purposes of this study, we classified funded PICU beds within each PICU as ‘Level 3’ if they were designated for children undergoing invasive mechanically ventilation outside of the operating room, emergency room or post-anesthetic care unit. We classified funded PICU beds as ‘Level 2’ if they were part of step down or high dependency units wherein invasive ventilation support is not offered.

### Participants

The internet-based survey (Appendix 1) was distributed to all 19 Canadian PICUs using a SurveyMonkey® link sent by members of the Pediatric Interest Group of the Canadian Critical Care Trials Group (CCCTG)(19). We excluded neonatal intensive care units that specialize exclusively in the treatment of premature babies and sick term neonates.

The survey was completed by a representative of each PICU in early 2022 and the collected data validated by PICU Directors in September-October 2022. Whenever clarifications about the collected responses were needed, the first author contacted the PICU directors and/or representatives. If the surveys were not completed within a week, the authors sent reminder emails or contacted the PICU representative via telephone.

### Analyses

Quantitative survey results were summarized as counts, proportions, and qualitative syntheses as appropriate. The funded beds in exclusively cardiac PICUs were included in the overall capacity analysis because these can be made available for general pediatric patients during surge situations with cancellation of elective cardiac surgeries. The following metrics were used to quantify the Canadian PICU capacity: number of funded and physical level 3 and 2 beds, provincial PICU bed density, number of physician Full Time Equivalents (FTE) per PICU and physician FTE-to-bed ratio. Although many Canadian PICUs admit patients up to the age of 18, we used the pediatric population (14 years or less) from the 2016 census data (https://www12.statcan.gc.ca/census-recensement/2016/dp-pd/index-eng.cfm) to calculate the PICU bed density (beds per 100,000 children) as this is the only category (0-14 years) provided in the Canadian census. For the calculation of PICU bed density, we combined the population of provinces/territories that did not have a PICU with that of provinces whose PICUs served them. We also calculated bed density for the entire population (beds per 100,000 people) using the Canadian census’ population data. Maps showing PICU sites and PICU bed density were generated using the BatchGeo™ (https://batchgeo.com/) platform and Microsoft Excel™ software, respectively. To calculate the physician-bed ratio, we divided the number of FTE positions per unit with the number of level 3 beds in the unit. We divided the annual number of admissions in individual units by the number of funded beds (level 3 + 2) to calculate admission rate per funded bed. For the free text portion of the survey related to challenges/ bottlenecks foreseen during surge situations, we used a general inductive content analysis approach (20). We read all the answers to a given question, keeping the objective in mind, generated initial categories inductively, then coded each response according to the developing framework, with categories added and adjusted as needed.

## Results

### Canadian PICU Capacity

We invited representatives from all **19** stand-alone PICUs in **17** hospitals across Canada and received a response from all units (100%) (figure 2). Within Canada there are **217** funded level 3 beds and **58** funded level 2 beds with a roster of **135.3** pediatric intense care physicians (Table 1). Together, these units reported having **298** physical bed spaces. Two PICUs representing **47** beds (**35** Level 3 and **12** Level 2) are specialized cardiac ICUs with a unique team of cardiac intensivists, nurses, and interprofessional clinicians that provide care exclusively for children with medical or surgical cardiac problems. Of the remaining units, 6 are mixed units, caring for children with cardiac and non-cardiac problems while 11 provide care for children with predominantly medical-surgical problems excluding cardiac surgical conditions.

**Table 1:** Canadian PICU Characteristics – Beds, physician staffing, services, isolation, and surge capacity

| PICU Location | Type | Funded Level 3 beds | Funded Level 2 beds | Funded Physician FTEs | ECMO availability | Surge beds | Isolation beds (%) | Retrieval team |
| --- | --- | --- | --- | --- | --- | --- | --- | --- |
| British Columbia |  |  |  |  |  |  |  |  |
| Vancouver | Combined | 12 | 0 | 10 | Yes | 12 | 76 - 100 | Yes |
| Victoria | Medical-Surgical | 5 | 0 | 2.75 | No | 3 | 26 - 50 | No |
| Alberta |  |  |  |  |  |  |  |  |
| Calgary | Medical-Surgical | 11 | 4 | 10 | Yes | 3 | 76 - 100 | Yes |
| Edmonton | Cardiac Surgical | 15 | 8 | 9 | Yes | 16 | 76 - 100 | Yes |
| Edmonton | Medical-Surgical | 15 | 12 | 9 | Yes | 10 | 76 - 100 | Yes |
| Saskatchewan |  |  |  |  |  |  |  |  |
| Saskatoon | Medical-Surgical | 12 | 0 | 5.5 | No | 6 | 26 - 50 | Yes |
| Manitoba |  |  |  |  |  |  |  |  |
| Winnipeg | Medical-Surgical | 9 | 0 | 5 | No | 6 | 76 - 100 | Yes |
| Ontario |  |  |  |  |  |  |  |  |
| London | Medical-Surgical | 12 | 2 | 6 | No | 4 | 51 - 75 | Yes |
| Hamilton | Medical-Surgical | 12 | 4 | 6 | No | 4 | 76 - 100 | Yes |
| Toronto | Medical-Surgical | 20 | 12 | 12 | Yes | 5 | 26 - 50 | Yes |
| Toronto | Cardiac Surgical | 20 | 0 | 8 | Yes | 0 | 26 - 50 | Yes |
| Kingston | Medical-Surgical | 0 | 4 | 1 | No | 6 | 76 - 100 | No |
| Ottawa | Combined | 10 | 6 | 7 | Yes | 5 | 76 - 100 | Yes (up to 3 y) |
| Quebec |  |  |  |  |  |  |  |  |
| Montreal Children's | Combined | 12 | 6 | 9 | Yes | 0 | 51 - 75 | No |
| CHU St. Justine | Combined | 24 | 0 | 14 | Yes | 14 | 76 - 100 | No |
| Quebec City | Combined | 12 | 0 | 9 | Yes | 4 | 0 - 25 | No |
| Sherbrooke | Medical-Surgical | 6 | 0 | 5 | No | 4 | 51 - 75 | Yes |
| Atlantic region |  |  |  |  |  |  |  |  |
| St. John's | Medical-Surgical | 4 | 0 | 3 | No | 4 | 26 - 50 | No |
| Halifax | Combined | 6 | 0 | 4 | Yes | 9 | 0 - 25 | Yes |
| TOTAL | -- | 217 | 58 | 135.25 | -- | 108 | -- | -- |

**Footnote:** PICU – Pediatric Intensive Care Unit; ECMO – Extra-Corporeal Membrane Oxygenation; FTE – Full-time Equivalent

**Figure 2:**
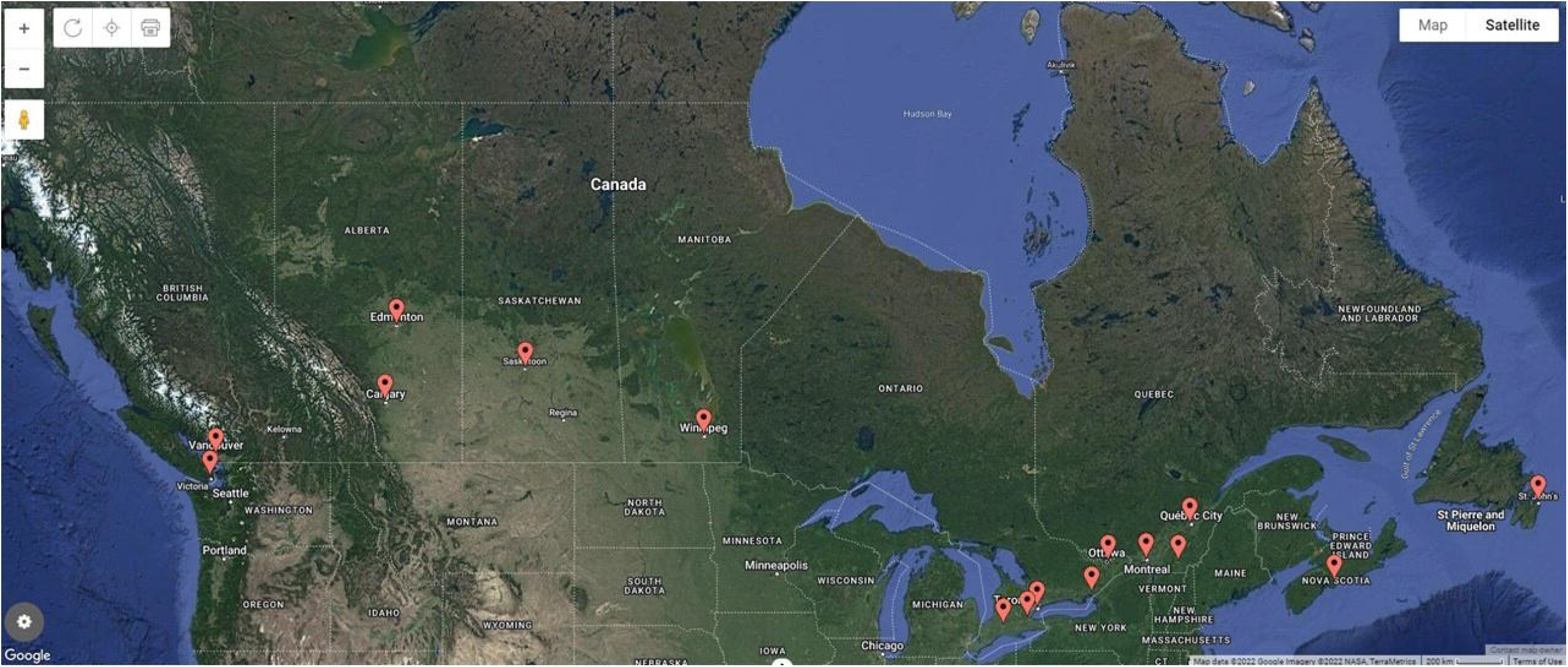
Map location of the 17 Canadian children’s hospitals housing the 19 Pediatric Intensive Care Units (created on *batchgeo*.*com®™*) Footnote: Montreal children’s hospital and CHU Sainte-Justine children’s hospital are located close to each other and are not distinguishable on this map.

Summarizing the distribution of PICU beds across provinces, there are no PICUs in Yukon, Nunavut, Northwest territories (NWT), Prince Edward Island (PEI) and New Brunswick (NB); these provinces are served by PICUs in BC Children’s Hospital (Yukon), Children’s Hospital of Eastern Ontario (Baffin Island, Nunavut), Alberta (NWT), and IWK Health (PEI and NB). The province with the highest PICU bed (medical-surgical & cardiac) density is Saskatchewan (5.56 beds/100,000 children 14-years or younger), while the provinces with the lowest PICU bed density are Nova Scotia, Prince Edward Island & New Brunswick (2.24 beds/100,000 children 14-years or younger children) (Figure 3).

**Figure 3:**
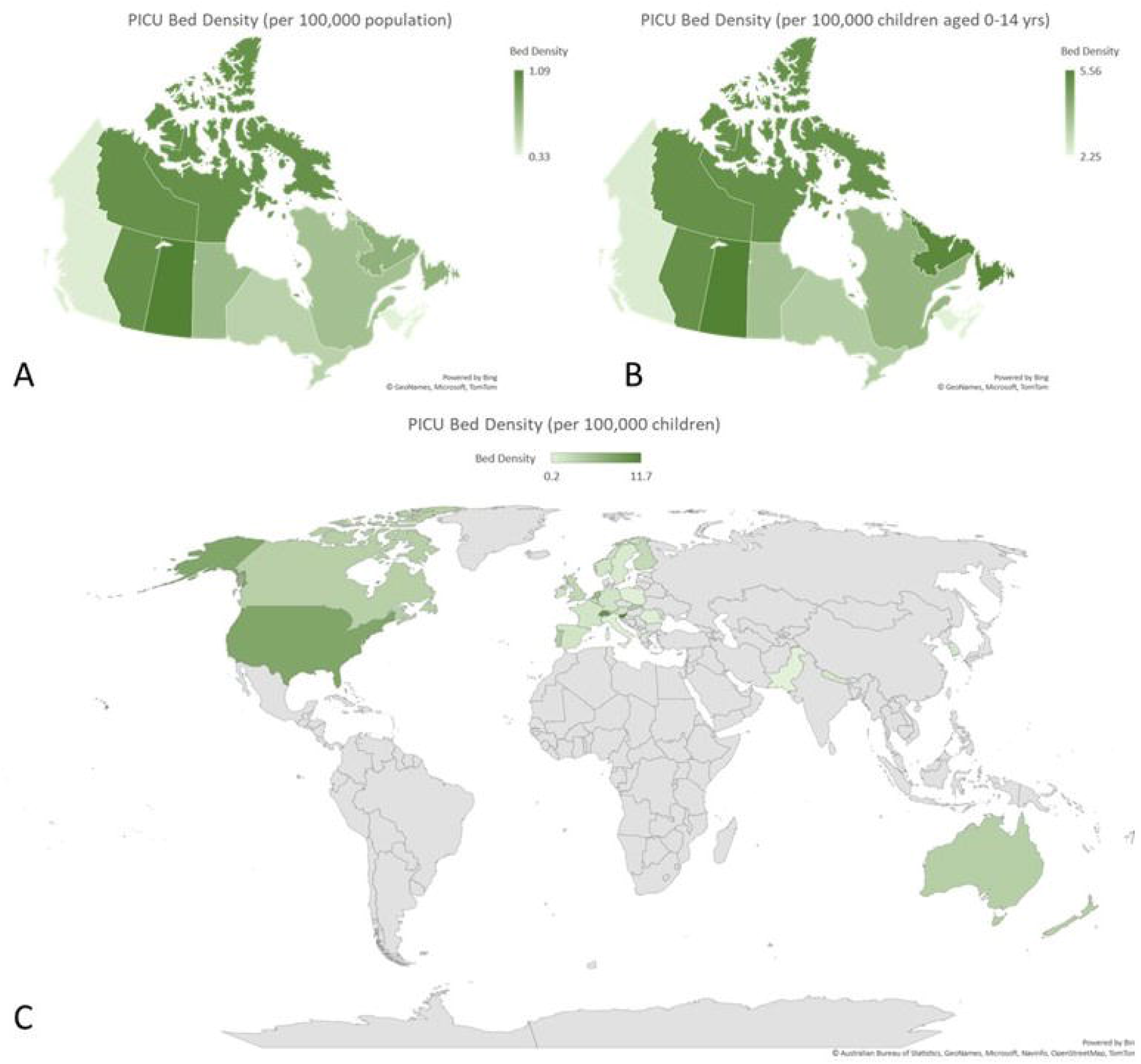
Pediatric intensive care unit bed density – Panel A: Canadian PICU bed density per 100,000 population); Panel B: Canadian PICU bed density per 100,000 children aged 0-14 years; Panel C: global PICU bed density per 100,000 children Footnote: • European data (survey yr. 2000) derived from Nipshagen et al. (27); Nepal data (yr. 2016) based on Khanal et al.(24); Pakistan data (survey yr. 2009) from Haque et al. (30); South Korea data (survey yr. 2015) based on Yoon JS et al. (34); United States of America data based on Horak RV et al. (26) • United Kingdom data from the PICANet report (32) (https://www.picanet.org.uk/wp-content/uploads/sites/25/2022/01/PICANet-2021-Annual-Report_v1.0-13Jan2022-2.pdf) • Canadian PICU bed density has been calculated based on population of 0-14-year-old children (not 0-18) • ANZ data based on that reported in the 2019-20-CCR-Activity-Report (28) (Source: https://www.anzics.com.au/wp-content/uploads/2021/06/2019_20-CCR-Activity-Report.pdf) • Nepal PICU bed density based on population of 27.26 million (2016) and children aged 0 – 14 yrs. old contributing to 32% of Nepalese population in 2016 (Source: https://www.statista.com/statistics/678090/nepal-children-as-a-percentage-of-the-population/)

A total of 135.3 full-time-equivalent (FTE) intensivist positions have been funded by the provinces and are currently filled by fellowship-trained pediatric critical care physicians. We also calculated the physician-per-funded level 3 bed and found that the mean ± SD value for Canada is 0.64 ± 0.14 (excluding Kingston, as Kingston did not report having any Level 3 beds).

### Population served

In the years 2018, 2019, 2020 and 2021, the 19 PICUs admitted roughly **13385, 13419, 11430 and 12315** Canadian children, respectively. Sherbrooke could only provide rough estimates (∼400 per year) regarding their annual admission number. Number of annual admissions and number of annual admissions per funded bed over these 4 years (2018 – 21) have been illustrated in figure 4a and 4b, respectively. Other details regarding these PICUs are provided in Table 1.

**Figure 4:**
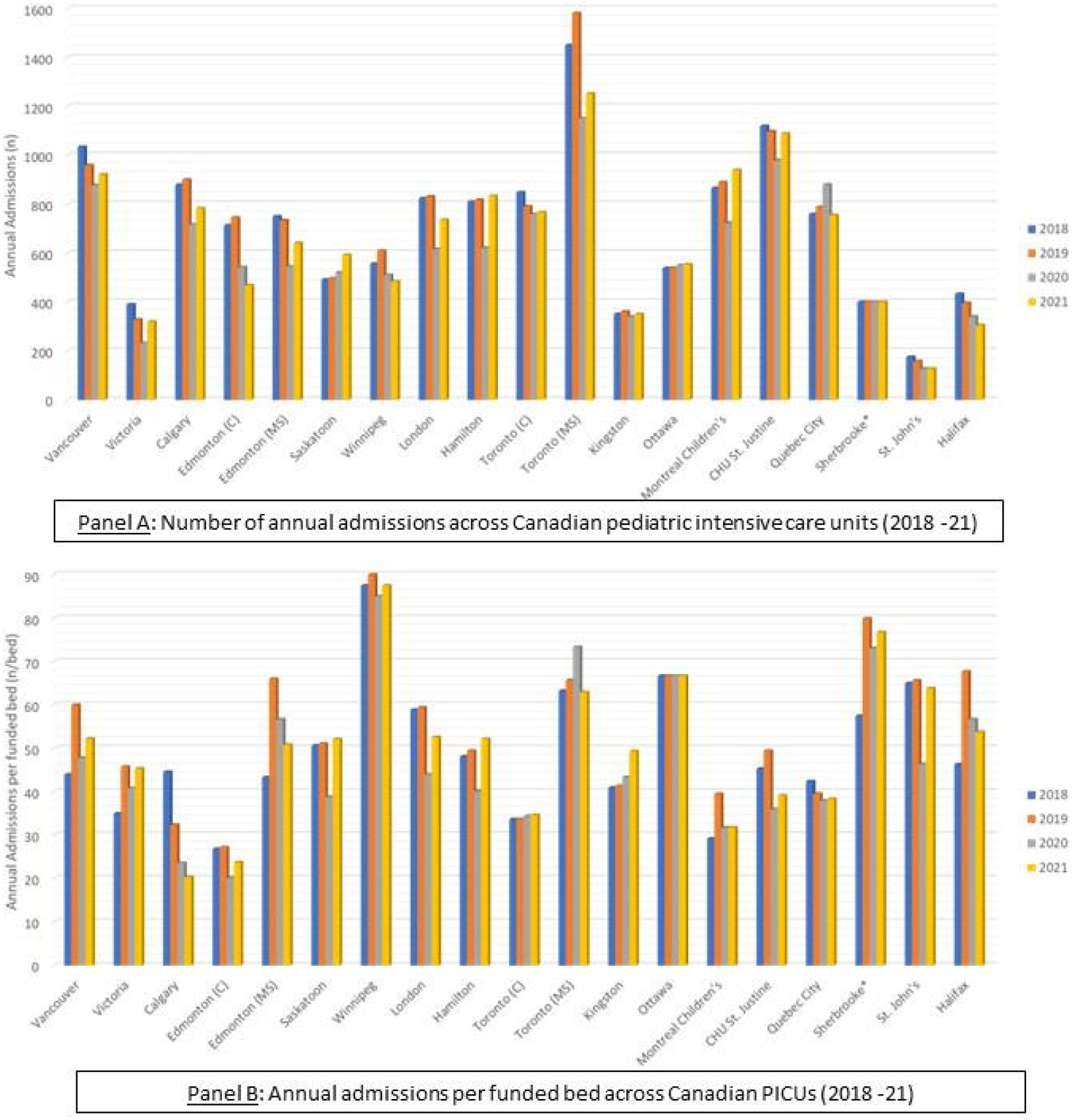
Number of annual admissions and annual admissions per funded bed across Canadian PICUs (2018 – 2021)

All 19 PICUs provide care to children aged 1 month to 16 years of age; 16 PICUs care for children up to 18 years old and 4 for young adults aged 19-20 years on a case-to-case basis. Neonates (0-28 days of life) are admitted in 18 out of 19 PICUs under the following circumstances: Neonate discharged home after birth and returns with critical illness (n= 10), neonates with congenital cardiac lesions (n=8), neonates requiring CRRT or ECMO (n=6), and neonates with surgical needs (e.g., congenital diaphragmatic hernia, bowel obstruction) (n= 3). One unit cares for critically ill peri-partum women. Seventeen PICUs provide care for children with polytrauma and/or neurosurgical problems, 16 for children with severe burns, and eight offer peri-operative care for solid organ transplant recipients.

### Services offered

In terms of other advanced monitoring or therapeutic services, 17 PICUs offer continuous electroencephalography (21). All 19 units reported that they have access to inhaled nitric oxide therapy and 18 offer high frequency oscillator ventilation (HFOV). Nine PICUs offer Extracorporeal Membrane Oxygenation (ECMO) and 18 offer continuous renal replacement therapy (CRRT). Of the 19 PICUs, 11 accept children transferred from other PICUs for advanced therapies. Both cardiac PICUs offer ventricular assistant device (VAD), a form of artificial heart support.

Children with infectious diseases that are airborne or spread through droplets require admission to a room with isolation capabilities. Two PICUs report that 0-25% of beds were isolation beds; four PICUs have 26-50% beds that can hold patients who require isolation; three PICUs stated that 51-75% beds had isolation capabilities while the remaining 10 units were able to hold patients requiring isolation in 76-100% of their beds (Table 1).

### Surge Management

All 19 PICUs had developed surge management plans. Our results showed that PICUs would be able to add **5.9 ± 3.4 (range: 0 – 14)** temporary surge beds. Canadian PICUs would be able to together create **108** additional temporary surge beds (Level 3 + Level 2). In a surge situation, respondents indicated that non-urgent cardiac and other surgical procedures would be ramped down or be completely halted to divert existing beds to care for critically ill children. That approach could potentially free up about 70% beds in cardiac surgical PICUs (approximately 32 beds) and 20% beds in medical-surgical PICUs (approximately 44 beds), accounting for the fact that emergency life-saving cardiac and non-cardiac surgeries would need to continue.

Several barriers for the successful implementation of surge plans were identified. We have summarized illustrative quotes, themes derived from the qualitative responses and emerging narratives in Table 3. Highlighted among these is the lack of situational awareness of PICU capacity and demands across the country to allow for more efficient inter-provincial collaborations.

**Table 2:** Pediatric critical care capacity estimates for Canada in comparison to other regions/countries

|  | Canada<br>(2021) | ANZ<br>(28)<br>(2020) | Europe<br>(27)<br>(2000) | USA<br>(26)<br>(2016) | United<br>Kingdom<br>(32)<br>(2020) | Spain<br>(29)<br>(1996) | Pakistan<br>(30)<br>(2009) | Nepal<br>(24)<br>(2016) | South<br>Korea<br>(34)<br>(2015) |
| --- | --- | --- | --- | --- | --- | --- | --- | --- | --- |
| Number of PICUs | 19 | 13 | 144 | 344 | 32 | 34 | 16 | 18 | 13 |
| Type | (n=19) |  | (n=104) |  |  | (n=31) |  | (n=16) |  |
| • Pediatric | 19 | 9 | 92 | 344 | -- | 18 | -- | -- | 12 |
| • Neonatal +<br>Pediatric | 0 | 1 | 8 | 0 | -- | 12 | -- | -- | 0 |
| • Adult +<br>Pediatric | 0 | 3 | 4 | 0 | -- | 1 | -- | -- | 1 |
| PICU subtype |  |  |  |  |  |  |  |  |  |
| • Medical-surgical | 11 | -- | -- | 285 | -- | -- | 12 | -- | 9 |
| • Cardiac | 2 | -- | -- | 49 | -- | -- | 3 | -- | 3 |
| • Mixed | 6 | -- | -- | -- | -- | -- | 1 | -- | 1 |
| • Trauma | 0 | -- | -- | 2 | -- | -- | -- | -- | -- |
| • Neuro | 0 | -- | -- | 8 | -- | -- | -- | -- | -- |
| PICU beds, n | 217 | 213** | 1198 | 5908 | 369 | 271 | 155 | 93 | 113 |
| PICUs with |  |  |  |  |  |  |  |  |  |
| • 6 or fewer<br>beds | 5 | -- | 21 | -- | -- | 15 | -- | -- | -- |
| • 7 – 12 beds | 7 | -- | 36 | -- | -- | 8 | -- | -- | -- |
| • 13 or more<br>beds | 7 | -- | 35 | -- | -- | 8 | -- | -- | -- |
| Funded/staffed<br>beds per PICU | 11.4 ±<br>5.9 | 11.5<sup>#</sup> | 9<br>(2 – 46) | 12<br>[8,20] | -- | -- | 9.7<br>(4 -28) | 5<br>[5-6] | 9.4<br>(2-30) |
| PICU beds per<br>100,000 children | 3.72* | 3.8 | 2.7 (0.5<br>-11.7) | 8 | 2.7(33) –<br>2.9^^ | -- | 0.2 | 1.1~ | 1.3 |
| Admissions per<br>year | 12,296<sup>@</sup> | 8,972 | -- | -- | 16,429 | 9,585 | 7,376 | -- | -- |

<sup>^</sup>PICANet report includes data from United Kingdom and Ireland (https://www.picanet.org.uk/wp-content/uploads/sites/25/2022/01/PICANet-2021-Annual-Report_v1.0-13Jan2022-2.pdf)
<sup>^^</sup> Based on national level population estimates by year, age and UK country -
https://stats.wales.gov.wales/catalogue/population-and-migration/population/estimates/nationallevelpopulationestimates-by-year-age-ukcountry
\* Calculated based on population of 0-14-year-old children
\*\* Physical beds as reported in the 2019-20-CCR-Activity-Report
<sup>#</sup> Based on available beds (Source: https://www.anzics.com.au/wp-content/uploads/2021/06/2019_20-CCR-Activity-Report.pdf)
<sup>~</sup>Calculated based on Nepal population of 27.26 million (2016) and children aged 0 – 14 yrs. old contributing to 32% of Nepalese population in 2016 (Source: https://www.statista.com/statistics/678090/nepal-children-as-a-percentage-of-the-population/)
<sup>@</sup>Sherbrooke provided approximate census data. Kingston does not have level 3 PICU beds (ventilated) and therefore their census was also not included.

**Table 3:** General inductive content analysis of challenges to mounting an effective surge response as identified by Canadian PICUs

| Category | Representative Quotes | Narrative description |
| --- | --- | --- |
| Shortage of human resources (n=17) | *"We have 6 physical beds, and, on most days, we only have 3-4 nurses on the schedule. Problems will arise if the skeleton nursing staff, we have become ill necessitating staying home. We will be relying on overtime and potentially nurses who no longer work in the PICU being re-deployed back"*<br>*"We have space and equipment to operate at 120-140% capacity but human resources will be the major issue."* | Lack of trained personnel, including critical care nurses, respiratory therapists & physicians to staff these additional or temporary surge beds could be the most important 'bottleneck' in mounting a surge response. |
| Limited number of beds (n=6) | *"We are often struggling to find a PICU bed when all our physical beds are occupied. We spend many hours on the phone trying to connect with other PICUs in the province, in the neighboring provinces and sometimes, even outside Canada to find a PICU bed for the critically ill child."*<br>*"Ward capacity is limited to accommodate PICU discharges and ED admissions."* | Shortage of beds in the PICU and the Pediatric wards keep some children waiting for a PICU bed and others in the PICU longer than necessary |
| Supporting adult patients (n=2) | *"There is also ongoing collaboration with the adult critical care such that there are adults being cared for in the pediatric ICU which is not yet set for an end date."* | A surge situation increasing critical care requirements for adults and children will redirect PICU resources to support adult patients |
| Staffing attrition (n=2) | *"Our RN staffing is much more challenged than previous due to burn-out, mat leaves, retirements, vaccine refusals and people deciding to leave the organization for various other reasons"* | PICU providers are experiencing an unprecedented attrition in numbers |
| Ramping down surgical procedures | *"Our surge plan involved our PACU, but this may not be feasible if we don't ramp down ORs like happened previously."* | Surgeries would have to be cancelled to allow PICUs to handle surge and this may not always be possible |
| Changing demographic of PICU admissions | *"Children with complex medical comorbidities requiring frequent PICU admissions seem to be growing in numbers."* | A higher proportion of critically ill children have medical comorbidities and have complex needs, and are staying in the PICU longer |
| Staffing models | *"Lack of governmental understanding that PICUs can't just ramp up their occupancy or the amount of work they do and looking at the average census doesn't allow us to staff for a surge."* | PICUs needs to reimagine their staffing models to be able to tackle surge situations |

## DISCUSSION

This study characterizes the national capacity and surge management capabilities of Canadian PICUs. Although this survey was carried out before the ongoing spike in critical care admissions due to viral respiratory infections in children across the country(13), results from this study are timely and are instrumental to plan for ongoing and future pediatric critical care surges. This work is also essential to raise awareness about the ongoing limitations in pediatric critical care capacity, as well as surge capacity, in Canada. It also provides data to inform future discussions among healthcare professionals and policymakers regarding the need for a coordinated national data platform to optimize pediatric critical care resources.

We found substantial variation in existing pediatric critical care capacity across regions, as well as varied ability to accommodate surges in pediatric critical care admissions across Canada. Such variation in current and surge capacity could result in differential decision-making about access to PICU care, and the services delivered during times of increased demand(5).

Like other countries, Canada’s PICUs are regionalized within larger urban areas (Table 2) (22–31). Our PICU bed-population ratio is comparable to that in Europe (yr. 2000) (27), United Kingdom(32, 33) and Korea (34). The USA is an outlier with its PICU bed density, comparable to the combined pediatric and adult ICU bed density in Canada (9.5 beds per 100,000)(5, 26). Canadian PICU bed density is comparable to that of USA’s Federal Emergency Management Area (FEMA) zone eight, which had the lowest PICU bed density of 5.6 per 100,000 children. While interpreting these ratios, we need to acknowledge that our definition of a PICU for this survey did not include adult intensive care units that occasionally admit/care for children, or Neonatal Intensive Care Units that intermittently in some regions do the same for children up to 2 years. Our assessment was also a cross-section of PICU beds and may not accurately represent the growth or trends in PICU capacity. In comparison to PICUs in USA (26), Canada does not have specialized PICUs solely focused on care of children with polytrauma or neurological problems.

It is important to contrast a) physical bed spaces that are not staffed with funded bed spaces that are resourced to provide high quality, safe pediatric critical care, and b) funded bed spaces that are yet to be operationalized with funded beds that are fully functional. For example, in response to the ongoing surge in acutely ill children with viral infections, provincial health authorities recently increased the number of funded PICU beds in some hospitals, but the physical bed spaces would need to be identified and the interprofessional team members to staff these newly funded beds would also have to be hired or reallocated from other hospital areas before these beds can become fully functional(35). For the purposes of our survey, we focused on funded PICU beds that have already been fully operationalized, and physical beds that have the space and necessary equipment to support a mechanically ventilated critically ill child but have not been staffed yet due to a lack of funding.

Although all Canadian PICUs reported having a crisis surge response plan, they may require further investment for updating and improvement. A sizable proportion of PICU beds are not designed to isolate patients with respiratory viral infections and may require redesigning of their physical layout for the safety of clinicians, patients, and families. Inability to implement appropriate infection control or isolation practices could lead to outbreaks and contamination of healthcare workers, further straining human resources during a surge and risking clinician stress and burnout(36, 37). Burnout among healthcare professionals, especially critical care nurses, has worsened since the onset of the COVID-19 pandemic as a result of, among other reasons, anxiety about risk of exposure, increasing demands for overtime, and sustained moral distress associated with caring for patients with COVID-19(38).

During the peak of COVID-19 waves 2 and 3, the Canadian adult critical care system was able to accommodate almost 150-200% of pre-pandemic averages by deferring elective surgeries/procedures, funding new ICU beds, identifying temporary surge spaces, redeploying staff (some from PICUs), and introducing team-based models of care (39). Current PICU surge management plans do not permit similar capacity. Furthermore, in contrast to other countries with nationally-coordinated registries of critically ill children (ANZICS – Australia & New Zealand Intensive Care Society(28) & PICANet – Paediatric Intensive Care Audit Network, United Kingdom(32)), data on PICU occupancy, diagnosis or case mix in Canada are only shared at a provincial level. Availability of real-time information regarding PICU bed availability, resource utilization, and de-identified data on patient case mix and outcomes would allow more effective, real-time decision-making and sharing of resources. Without a coordinated central database, PICUs in Canada will continue working in isolation, lacking coordination and situational awareness to respond in times of crisis.

Our study has important limitations. First, as a cross-sectional survey of Canadian PICU capacity we cannot account for the possibility of increased or reallocated resources over time. There may have been changes, either temporary or permanent, to the number of PICU beds in some centers in response to the recent surge and our study does not account for such changes. However, we were careful in requesting such information and we believe that our surge capacity was well reported by the respondents across all PICUs in the country before the fall 2022 surge. Second, reporting of capacity was done by one representative and validated by the clinical director from each PICU. Though we carefully requested exact numbers and checks, reported and actual practice may not be the same(19). Third, we did not explore the PICUs’ daily census/occupancy trends, which are valuable but harder to collect as part of a survey. Studies show that ICUs that consistently operate in a high occupancy state (85-100%) have a 19% (0 – 44%) higher adjusted patient mortality risk(40). Lastly, the PICU census is known to have a significant seasonal variation with peaks during winter months (41, 42). Although our study did not explore the seasonal variations in PICU census, PICU bed funding as well as allocation of bedside staff must be planned so that patient care does not suffer, and staff are not subjected to unsafe working environments during periods of predictable or unpredictable high patient census. Therefore, future studies should explore day-to-day census and operational aspects of Canadian PICUs, as well as the ability to respond to surges by quickly providing critical care training to nurses, respiratory therapists, and other members of the inter-professional team.

## Conclusion

Canadian pediatric critical care capacity is comparable to other high-income countries, though response to an eventual pandemic/epidemic surge with significant pediatric critical illness may be limited. Federal and provincial governments should collaboratively and proactively plan sustainable increases in long-term Canadian PICU capacity including provision for centralized data coordination and improved patient isolation.

## Data Availability

All data produced in the present study are available upon reasonable request to the authors.

